# Bridging machine learning and compartment models to predict an epidemic

**DOI:** 10.1101/2022.10.07.22280853

**Authors:** Minh Triet Chau

## Abstract

This work proposes a Physics-informed Machine learning method to model and emulate the progression of COVID-19. Besides the high accuracy, lower data need, and interpretability, the method also estimates hidden parameters from data, which are useful for policymakers to flatten the curve and better understand public healthcare system.

## 1 Introduction

It is beneficial to forecast the course of the pandemic to plan effective control strategies. There are two approaches to this prediction problem, the first is purely data-driven techniques without epidemiology prior knowledge [Alfred and Obit, 2021]. On the other hand, epidemiologists develop compartment models that embed their prior knowledge [William and Anderson, 1927, Aron and Schwartz, 1984]. As the COVID-19 epidemic poses more questions than these models can answer, scientists develop more sophisticated models with more parameters, but it is getting more complicated to tune them manually. We propose the use of machine learning (ML) to optimize the parameters fitting a compartment model [Giordano et al., 2020] and compare it with data-driven methods. The source code is available at https://github.com/minhtriet/covid_ode.

This work uses the Canada data from [Dong et al., 2020]. It contains the number of infected, recovered, and death cases. Due to quality control difficulties, they discontinue tracking the number of recovered data after 4th August 2021^1^. To better demonstrate the selected compartment model, this work only uses the time frame when the numbers of infected, recovered, and deaths are available, which contains 555 days. We split the data into train, validation, and test sets whose lengths are 277, 55, and 223 days, respectively. A visualization is in Figure 1.

**Figure 1:**
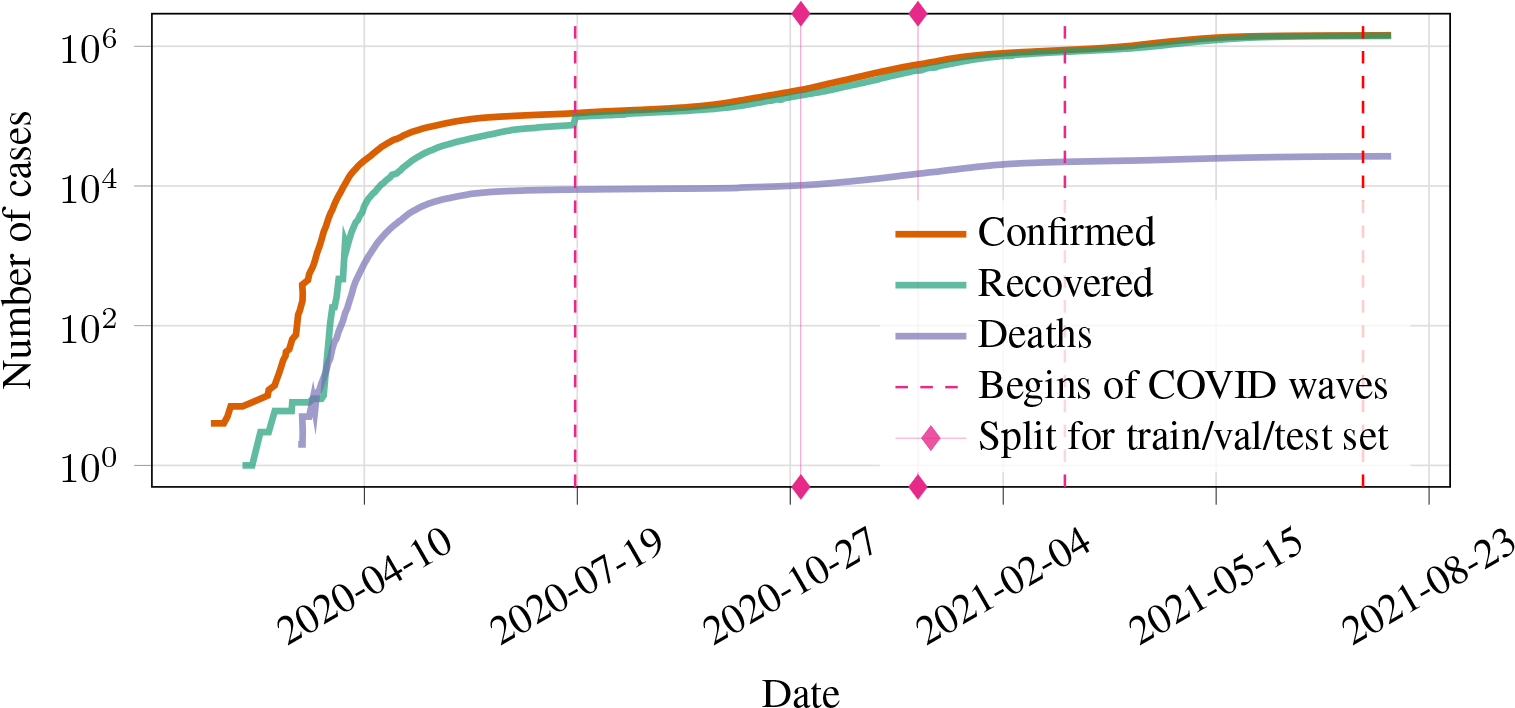
COVID in Canada through figures. The number of cases is on the log scale. The days of the beginning of the waves are from [Public Health Agency of Canada, 2022]. There is one infection wave (the dashed line) in the training set, zero waves in the validation set, and two waves in the test set.

## 2 Methodology

We experiment with ARIMA, Temporal Fusion Transformers (TFT) [Lim et al., 2021], and our proposed method. ARIMA is an earlier autoregressive model integrated with the moving averages. On the other hand, TFT is a more modern DL method that uses attention mechanism [Vaswani et al., 2017]. Attention mechanism achieved impressive result in Natural language processing, as is widely adapted to time series forecast. ARIMA and TFT implementation is from [Pedregosa et al., 2011] and [Herzen et al., 2022], respectively. A shared property of all three architectures is their internal parameters that we have to fit with the training data.

Designed specifically to model COVID-19, [Giordano et al., 2020] partition the population into eight classes (Table 1a) with transition probability parameters between them (Table 1b). Unlike the other compartment models, it discriminates between detected and undetected cases of infection and between different severity of illness. This separation helps policymakers estimate the number of unknown patients hidden in the community.

**Table 1:**
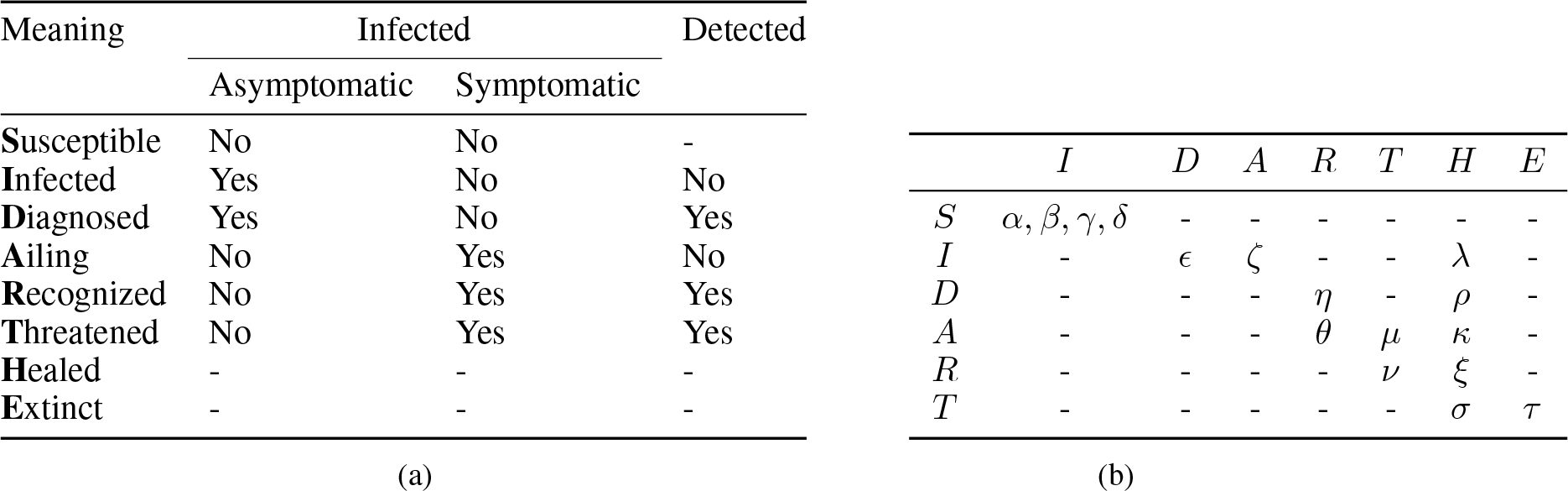
Terms and their explanation for the SIDARTHE model. In Table 1a, **T** means that the patient has life threatening symptoms while **R** does not. In Table 1b, the parameters map terms from the row to the column. While the rest of the parameters are self-explanatory, the meaning of *α, β, γ*, and *δ* are more specific. Concretely, *α* is the transmission rate of an *S* from *I, β* from *D, γ* from *A*, and *δ* from *R*.

When feeding the input data to this model, it is unclear whether to map the number of infected people to *R* or *D*, as the number reported in the data does not differentiate between asymptomatic and symptomatic infected. We settled on mapping it to *D* and using the parameter *η* as the transition from *D* to *R*. Besides that, *H* and *E* map directly to the number of recovered and death present in input data. All of the other parameters are unknown and learnable.

The training procedures follow [Wang et al., 2021]. We first use the Runge–Kutta 4 implementation of [Chen, 2018] to discretize and approximate from the current model’s parameters. After that, we compute the loss function based on *D, H, E* and backpropagate the error to tune every parameter until convergence.

## 3 Experiment

The goal is to minimize a loss function *L* between the predicted and actual results. In the case of root mean squared error loss (RMSE), 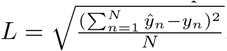, with *N* the number of data points, *ŷ, y*_*n*_ the prediction and actual data. To be closer to reality, in the test phase, we use its past data of the test phase to continuously retrain all of the models.

### 3.1 Baseline: ARIMA and TFT

Since DL methods depend on the hyper-parameters they are initialized with, in the case of TFT, we perform a grid search to get the best values for the number of encoder and decoder layers, dropout rate, number of input days, and the number of attention heads. TFT would take *I, E*, and *R* as a a multivariate time series, while ARIMA only supports univariate time series. We also use the number of input dates used for TFT for ARIMA.

Even though TFT reports a low training error, it fails to extrapolate. It is not unexpected, as TFT has only hundreds of training data points to tune a large number of parameters (around 21000). Therefore it is prone to overfitting. Retraining during the test phase does not help improve the performance of TFT. Regardless of their performance, however, the values of parameters of both TFT and ARIMA do not help in terms of epidemiology and control. We discuss a model that addresses this shortcoming in the next session.

### 3.2 ML to train the compartment model

The compartment model takes the shortest time to train and has the best performance compared to other experimented methods (See Table 3). Therefore, we assume that the values of the hidden parameters after training fit the data well. As each parameter either helps gauge the healthcare system or corresponds to a different policy, we discuss more of them in Table 2. The RMSEs are in 3 and the visualization of the prediction is in Figure 2.

**Table 2:** The values of hidden parameters and discussion. Table 1b contains more information about the above parameters.

| Parameter | Value | Discussion |
| --- | --- | --- |
| *Transmission rate* |  |  |
| $\alpha$ | 0.4497 | $\alpha > \gamma$ means people avoid individuals with symptoms. $\gamma > \beta, \delta$ (assuming that subjects who have been diagnosed are properly isolated). These parameters correlate social-distancing or face masks mandate |
| $\beta$ | 0.3382 | |
| $\gamma$ | 0.4148 | |
| $\delta$ | 0.1402 | |
| *Detection rate* |  |  |
| $\epsilon$ | 0.1708 | $\theta > \epsilon$ means a symptomatic subject is more likely to be tested than an asymptomatic. |
| $\theta$ | 0.4218 | |
| *Develop relevant symptoms rate* |  |  |
| $\zeta$ | 0.0863 | These parameters correlate with the community adaptive immunity [Boyton and Altmann, 2021] |
| $\eta$ | 0.0100 | |
| *Develop life-threatening symptoms rate* |  |  |
| $\mu$ | 0.1974 | $\mu > \nu$ means undetected cases are more likely to turn critical than detected cases. |
| $\nu$ | 0.0100 | |
| *Mortality rate* |  |  |
| $\tau$ | 0.1013 | This is higher than reported by [Government of Canada, 2022] (approximately 0.0007% around the test time). However, note that it is the death rate from $T$. |
| *Recovery rate* |  |  |
| $\lambda, \kappa, \xi,$<br>$\rho, \sigma$ | 0.0100 | The higher these parameters are, the better the treatments and community immunity |

**Table 3:** The RMSE of ARIMA, TFT, and our approach.

| Input | ARIMA | TFT | Our method |
| --- | --- | --- | --- |
| Confirmed | 32189.884 | 46690140.864 | **16410.667** |
| Recovered | 28208.252 | 41675153.819 | **10267.877** |
| Death | 809.910 | 2958386.149 | **168.567** |

**Figure 2:**
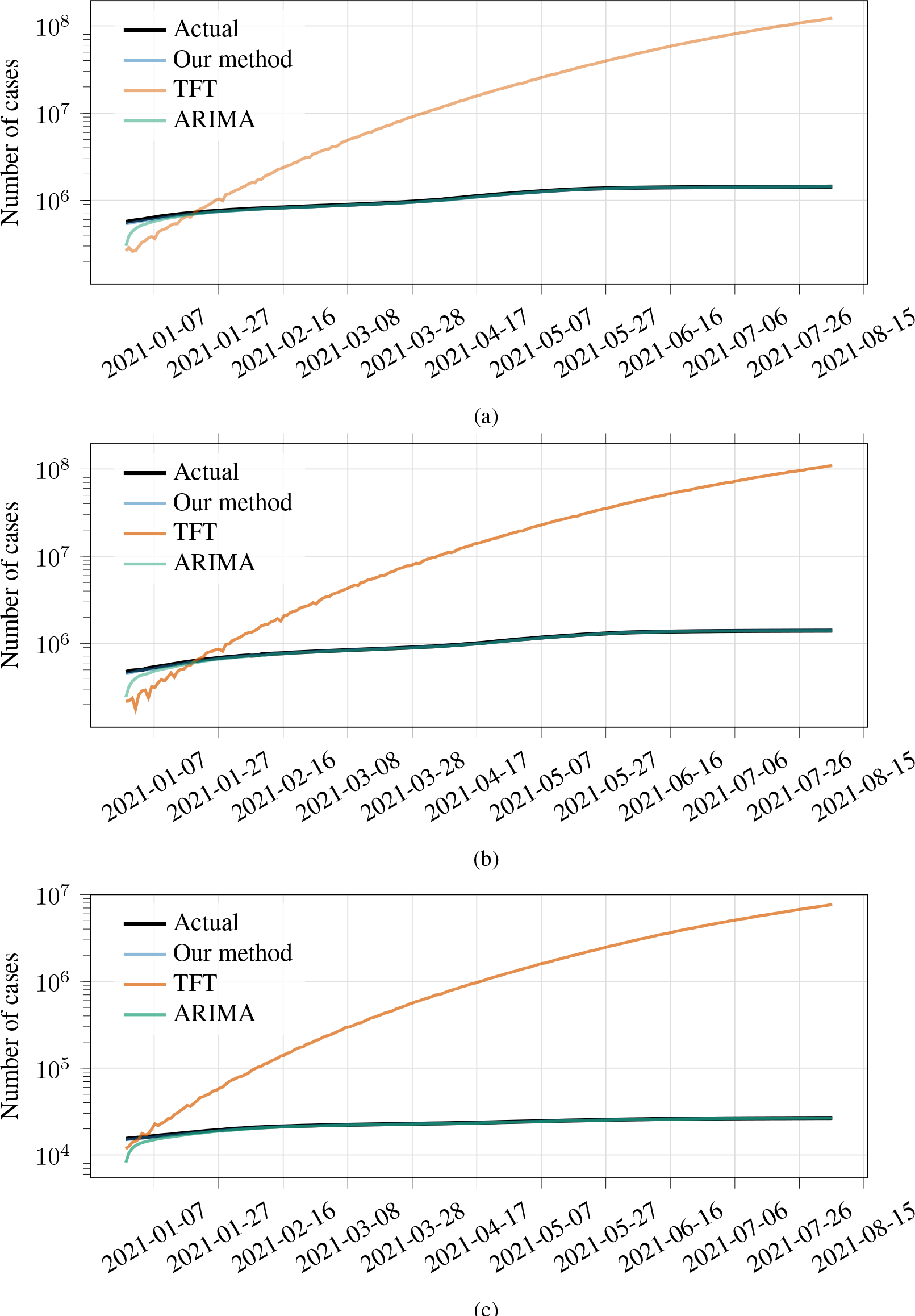
The prediction performance for each quantity of each method. Figure 2a, 2b, and 2c show the prediction of each method for the number of infected, recovered, and deaths, respectively. TFT consistently overestimates the number of cases, while our method and ARIMA have much smaller error margins.

## 4 Conclusion

We pose COVID-19 as a non-linear dynamical system and propose the use of ML and a compartment model for forecasting. DL suffers from the low number of data samples and the distribution shift in the epidemic dynamics. On the contrary, through different infection waves, the ML model was able to learn sensible values to the parameters and produced an accurate prediction. Its parameters for unobservable data are important to understand the course of the epidemic and keep track of the healthcare system.

## Data Availability

All data produced in the present study are available upon reasonable request to the authors

## Footnotes

1 https://github.com/CSSEGISandData/COVID-19/issues/4465

